# Triangulating Evidence for Antihypertensive Drug Class Efficacy on Cardiovascular and Metabolic Outcomes Using Mendelian Randomisation and Colocalisation

**DOI:** 10.1101/2024.08.22.24312458

**Authors:** Nhu Ngoc Le, Tran Quoc Bao Tran, John McClure, Dipender Gill, Sandosh Padmanabhan

## Abstract

**Background:** Current hypertension treatment guidelines typically recommend a standardised approach, which may not account for the inter-individual variability in blood pressure (BP) response or the complex causation of hypertension. This study aims to investigate the heterogeneity of responses to a broad range of antihypertensive drugs across various cardiometabolic and renal outcomes.

**Methods:** This study employed an integrative approach combining Mendelian randomisation (MR), summary-based MR (SMR), and colocalisation analyses to investigate the impact of BP lowering and the efficacy of seventeen antihypertensive drug classes on the risk of coronary artery disease (CAD), myocardial infarction (MI), atrial fibrillation (AF), heart failure (HF), ischemic stroke, chronic kidney disease (CKD), and type 2 diabetes (T2D). Genetic association and gene expression summary data were obtained from the largest European ancestry GWAS and GTEx v8 for 29 tissues that were broadly relevant to the pathophysiology of cardiovascular outcomes included.

**Results:** The genetic evidence supported that lower SBP was universally beneficial, causally associated with reduced risks of all studied outcomes. The association of genetically predicted SBP lowering varied significantly depending on the antihypertensive drug class, revealing heterogeneity in their impact on different health outcomes. Novel MR associations were identified, including protective effects of endothelin receptor antagonists, sGC stimulators, and PDE5 inhibitors against CAD (per 10-mmHg decrease in SBP, OR range = 0.197 - 0.348) and ischemic stroke (OR range = 0.218 - 0.686); and sGC stimulators and PDE5 inhibitors against CKD risk (OR range = 0.532 - 0.55). SMR and colocalisation analyses include evidence for *GUCY1A3* and CAD and MI risk, *KCNH2* with AF risk, and *PDE5A* with CAD risk.

**Conclusions:** Our results support potential differential impacts of antihypertensive drug classes on cardiometabolic and renal outcomes, underscoring the potential for personalised therapy. Future research should validate these findings across diverse populations and explore the mechanistic pathways between antihypertensive BP modulation and health outcomes.

**Clinical Perspective:** *What is New?:*

- This study utilised Mendelian randomisation, summary-based MR, and colocalisation analyses to explore the differential effects of 17 antihypertensive drug classes on various cardiometabolic and renal outcomes.
- Although lower SBP is universally beneficial for reducing risks across all outcomes, the effectiveness of SBP lowering varies significantly by antihypertensive drug class, showing heterogeneity in their impact on different health outcomes.
- Newer therapies, including ERAs, PDE5 inhibitors, and sGC stimulators, showed significant protective effects across various outcomes, with ERAs benefiting AF, CAD, MI, and stroke; PDE5 inhibitors protecting against all outcomes except T2D; and sGC stimulators being effective against CAD, MI, stroke, T2D, and CKD.

*What are the Clinical Implications?:*

- Understanding the varying impacts of different antihypertensive drug classes on health outcomes can guide more personalised treatment strategies, potentially improving patient outcomes.
- These results not only validate the clinical relevance of existing antihypertensive therapies but also highlight new therapeutic targets, such as *GUCY1A3* and *PDE5A*, for further exploration.
- Future research should focus on validating these results across diverse populations to refine hypertension management strategies and implement personalised treatment in clinical practice.

## Introduction

Blood pressure (BP) regulation is a complex, multifactorial process influenced by a range of genetic, environmental, and lifestyle factors (1). Hypertension, defined as a BP level above a specific threshold associated with an increased risk of cardiovascular outcomes, is similarly multifactorial (2). Current hypertension treatment guidelines generally recommend an empirical approach, where the initial treatment involves the use of one or two first-line antihypertensive agents from different pharmacological classes based on the severity of BP elevation (3–5). If BP remains uncontrolled, additional agents are sequentially introduced. Given the complexity of hypertension pathophysiology, it remains unclear whether the standardised approach to treatment is the optimal strategy for BP control. The question of whether individually targeted BP treatments can maximise clinical benefits has not been thoroughly explored.

Randomised clinical trials (RCTs) have established the efficacy of commonly used antihypertensive drugs (6, 7), leading to the perception that the case for personalising therapy is more academic than clinically relevant. Nevertheless, despite regularly updated guidelines and a wealth of clinical evidence, a significant percentage of adults with hypertension continue to have uncontrolled BP despite ongoing antihypertensive therapy (8). This persistent issue suggests that a re-evaluation of treatment strategies may be necessary.

There is substantial evidence for intra- and inter-individual variability in BP response to different antihypertensive drugs partially explained by factors such as genetics, age, sex, dietary factors and baseline BP levels (1). Additionally, variability in long-term outcomes has been observed depending on the antihypertensive drug class; for example, ACE inhibitors and ARBs have demonstrated superior efficacy in reducing the risks of heart failure and chronic kidney disease (9, 10). The recent PHYSIC trial, employing a randomized repeated cross-over design, further highlighted significant variability in individual BP responses to four commonly used antihypertensive drugs, suggesting that personalised therapy could achieve greater BP reductions (11). However, evidence gaps remain, particularly concerning other antihypertensive drug classes and a broader range of clinically important outcomes, such as coronary artery disease (CAD), stroke, heart failure (HF), atrial fibrillation (AF), chronic kidney disease (CKD), and type 2 diabetes (T2D). Addressing these gaps through randomized controlled trials (RCTs) is likely to be both challenging and impractical.

Over the last 15 years, genome-wide association studies (GWAS) have provided a wealth of genetic data, enabling novel analytical approaches to study the causal effects of risk factors and therapeutic interventions on health outcomes (12).

Mendelian randomisation (MR) (13), summary-based Mendelian Randomisation (SMR) (14), and colocalisation analyses (15) are powerful tools that leverage genetic variants as instrumental variables to infer causality and elucidate the mechanisms underlying complex traits. By using genetic proxies for BP, MR can elucidate the long-term effects of BP-reducing drug targets on various health outcomes, relatively devoid of confounding factors and reverse causation (13). SMR further extends this approach to identify causal effects of gene expression on traits, while colocalisation distinguishes between pleiotropy and shared genetic causation (14, 15).

In this study, we propose to explore the heterogeneity of responses to a majority of antihypertensive drugs across a wide range of cardiometabolic and renal outcomes through an integrative approach combining MR, SMR, and colocalisation analyses.

By combining multiple methods along with triangulation of evidence, this study seeks to strengthen the case for personalised therapy for hypertension, challenging the standardised treatment approach currently advocated in clinical guidelines.

## Methods

Study design: Our study includes two stages: (i) perform two-sample Mendelian randomisation (MR) to explore potential causal effects of SBP reduction and antihypertensive drug classes on a range of cardiovascular, diabetic, and renal outcomes; (ii) perform summary-based MR and colocalisation to examine if any detected drug-disease association is mediated through gene transcription, integrating summary data from GWAS and expression quantitative trait loci (eQTL) studies (**Figure 1**).

**Figure 1.**
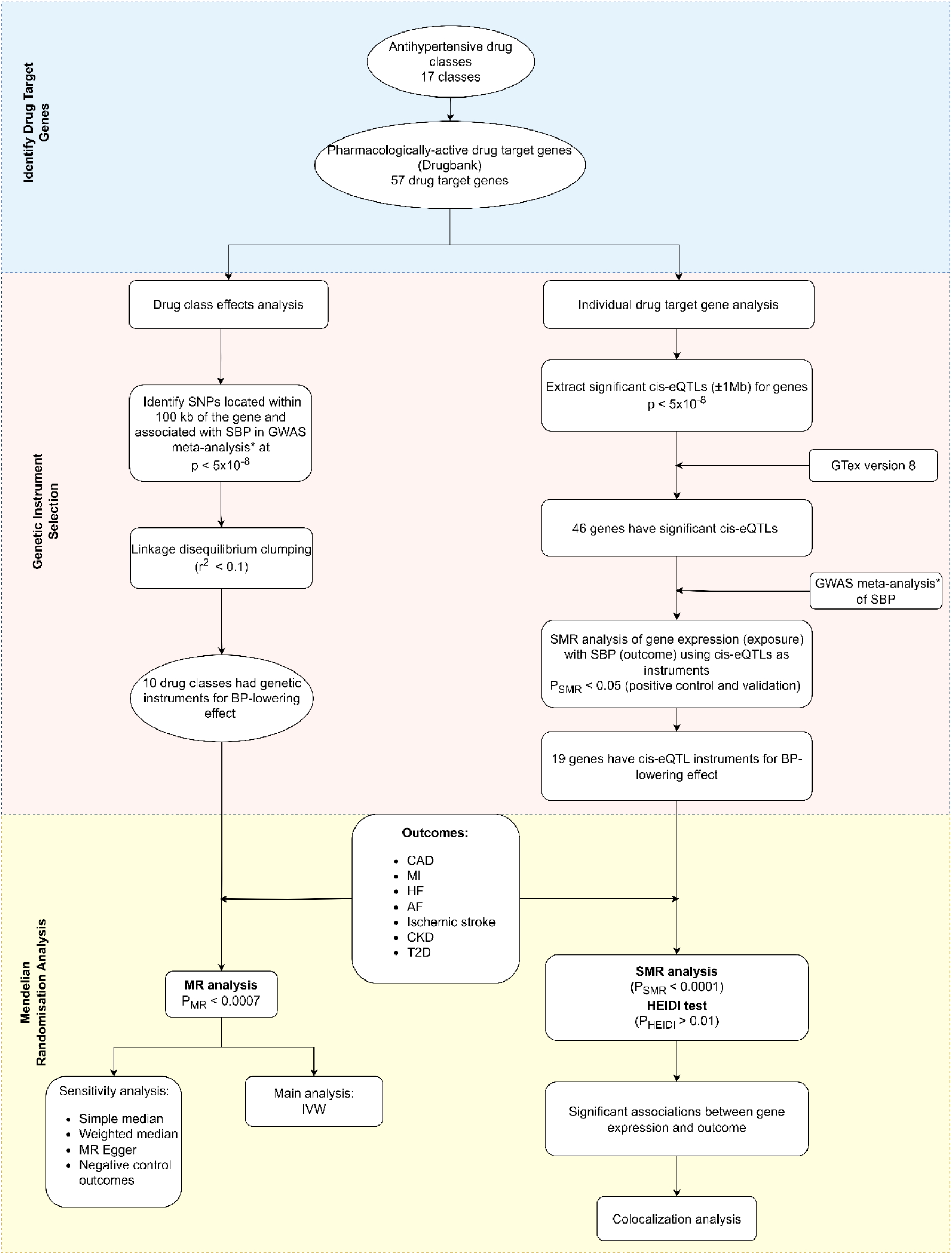
Study design. SBP, systolic blood pressure; GWAS, genome-wide association study; SNP, single nucleotide polymorphism; eQTL, expression quantitative loci; MR, Mendelian randomization; CAD, coronary artery disease; MI, myocardial infarction; AF, atrial fibrillation; HF, heart failure; CKD, chronic kidney disease; T2D, type 2 diabetes; IVW, inverse variant weighted; (*) indicates a blood pressure GWAS meta-analysis (18).

All data used in this study are publicly available summary data extracted from GWAS and eQTL studies, with ethical approval and informed consent from all participants being obtained from their original studies.

### Identification of antihypertensive drug classes and their drug target genes

17 antihypertensive drug classes were identified by reviewing the British National Formulary (BNF) (16). These are angiotensin-converting enzyme inhibitors (ACEi), angiotensin II receptor blockers (ARBs), calcium channel blockers (CCBs), beta-adrenoceptor blockers (BBs), thiazides and related diuretics (TZDs), alpha-adrenoceptor blockers, adrenergic neuron blocking drugs, centrally acting antihypertensive drugs, loop diuretics, potassium-sparing diuretics (PSDs), aldosterone antagonists, renin inhibitors, endothelin receptor antagonists (ERAs), phosphodiesterase type 5 (PDE5) inhibitors, soluble guanylate cyclase (sGC) stimulators, and vasodilator antihypertensives. An emerging therapy included in our study is Zilebesiran (17), an investigational RNA interference agent that inhibits the production of hepatic angiotensinogen, which has shown effectiveness in lowering BP (**Table S1**).

Pharmacologically active protein targets of each drug class and the corresponding genes were identified using the Drugbank database. In total, we obtained 57 drug target genes (**Table S1)**.

### Selection of genetic instruments for MR analysis of drug class effects

To proxy the effect of antihypertensive drug target perturbation, genetic instruments were selected as single nucleotide polymorphisms (SNPs) located within 100 kb on either side of each drug target gene and strongly associated with systolic blood pressure (SBP) at a genome-wide significant level p-value < 5×10^-8^. Summary statistics for SBP were obtained from a GWAS meta-analysis of 757,601 European-ancestry participants drawn from the UK Biobank and the International Consortium of Blood Pressure (ICBP) (18).

For some drug classes whose protein targets are encoded by different genes, genetic instruments for each gene were combined as a single set of instruments for the drug effect analysis (19).

Linkage disequilibrium (LD) clumping at r^2^ < 0.1 was conducted to remove highly correlated SNPs. F-statistics were calculated to evaluate the strength of individual genetic instruments (20).

### Selection of genetic instruments for MR analysis of SBP-lowering effects

SNPs associated with SBP at a p-value < 5×10^-8^ (18) across the whole genome were selected as genetic instruments for SBP. Due to the large number of SNPs identified, a more stringent threshold r^2^ < 0.001 was used for the clumping to minimise potential bias from LD. The instrument strength was quantified using R^2^ and F-statistics (20).

### Selection of genetic instruments for summary-based MR analysis of drug target gene expression

Cis-expression quantitative loci (cis-eQTL) were selected as genetic instruments to proxy for the effect of changes in gene expression level. Only cis-eQTL located within 1 Mb window of the gene were considered for the selection. Significant cis-eQTL data (p < 5×10^-8^) were obtained from GTEx version 8 (49 tissues, from nearly 1000 deceased individuals, predominantly European ancestry) (21). The eQTL data is on a scale of 1 unit change in gene expression level per each additional effect allele.

For positive control testing and validation, the association of gene expression with SBP were examined - only genes whose expression levels were associated with SBP (p < 0.05) were taken forward for the SMR analysis on the outcomes.

### Outcome data

Outcomes under this study include CAD, MI, AF, HF, ischemic stroke, CKD and T2D. These outcomes were selected for the study based on their clinical significance and relevance to hypertension and BP-lowering drug effects (22–25). GWAS summary data for these outcomes were obtained from the largest GWAS available, with most participants of European ancestry to align with our exposure data (**Table 1**).

**Table 1.**
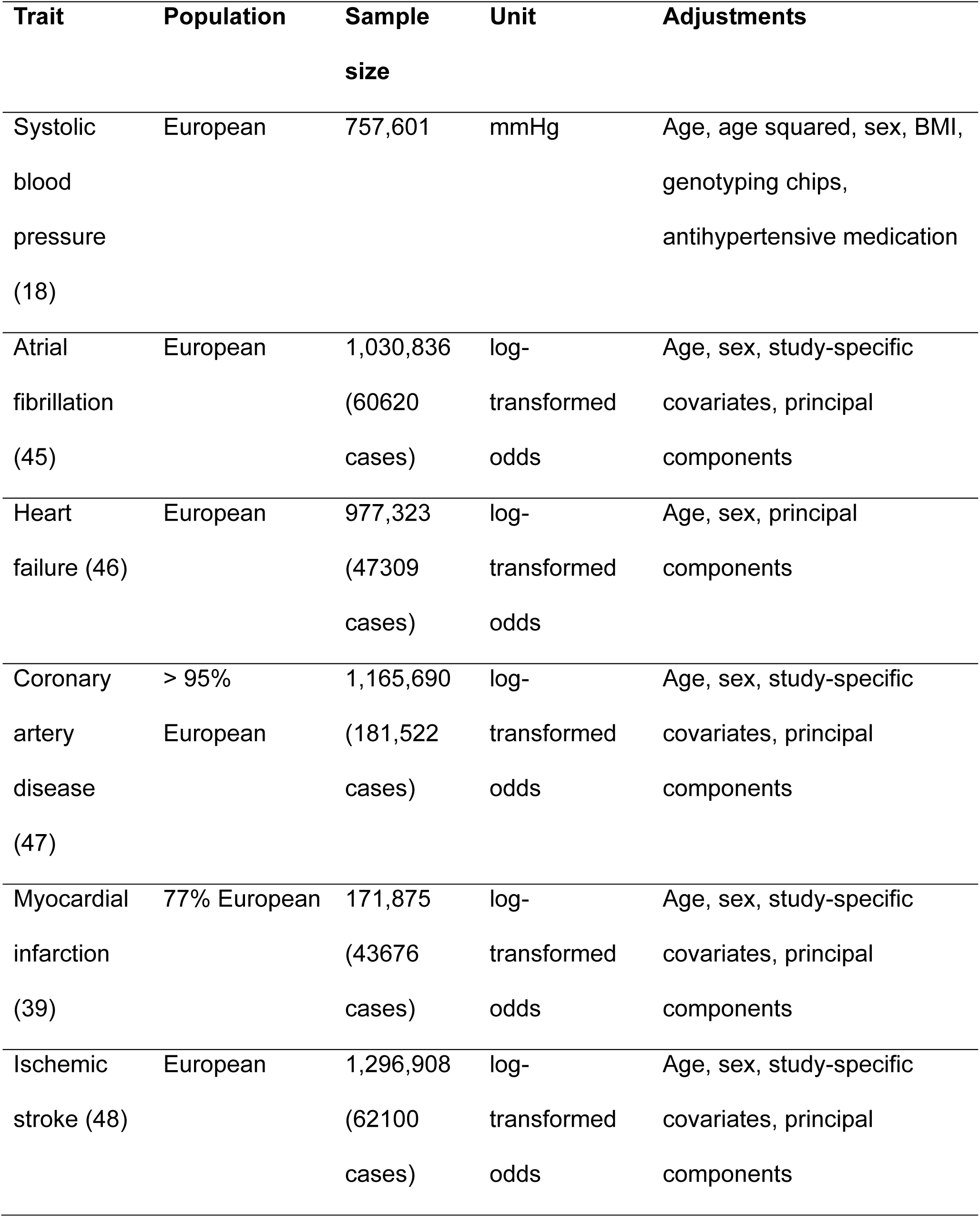
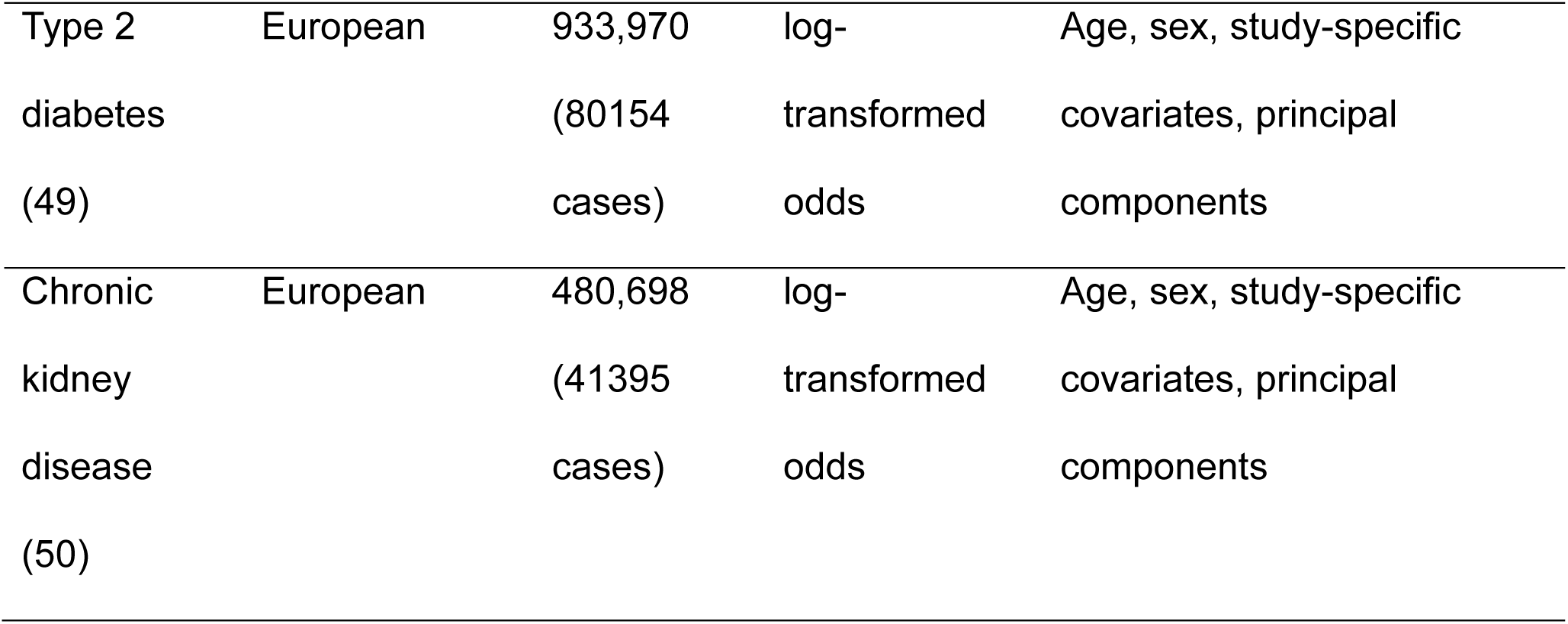
GWAS used in MR analyses.

### Two-sample Mendelian randomisation analysis

MR is a form of instrumental variable analysis, which uses genetic variants as proxies to evaluate the potential causal effect of a modifiable exposure on health outcomes. MR relies on three main assumptions: (i) the genetic variant is strongly associated with the exposure; (ii) the genetic variant is not associated with any confounders relating to the association between the exposure and the outcome; (iii) the genetic variant is not associated with the outcome, except through the exposure.

In this two-sample MR analysis, genetic variants located in the vicinity of a gene that encodes the BP-lowering drug target were used as instruments to investigate the causal relationship between antihypertensive drugs and the outcomes. Data harmonisation was performed to ensure the variant-exposure association and the variant-outcome association are aligned to the same effect allele. Palindromic variants were not excluded from the analysis. To ensure consistency in genetic variants used across MR analyses, proxies were not used in our study.

The inverse-variant weighted (IVW) method was used as the main MR method. MR estimates were presented as odds ratio (OR) for each outcome, with a 95% confidence interval (CI) per 10 mmHg decrease in SBP. This is to reflect the BP-lowering effect of antihypertensive drug classes. To account for multiple testing, a Bonferroni-corrected p-value of 0.0007 (0.05 / 11 (drug classes & SBP) × 7 outcomes) was used to detect significant associations; those with a p-value greater than 0.0007 but less than 0.05 were considered nominal associations.

The simple median (26), weighted median (26) and MR Egger (27) methods were performed as sensitivity analyses to evaluate the robustness of MR findings to potential bias. These methods have less stringent assumptions compared to the IVW method. Simple median and weight median methods require at least 50% of the genetic variants to be valid to give a robust MR estimate. MR Egger can give a consistent MR estimate of the causal effect when up to 100% of genetic variants are invalid but require the Instrument Strength Independent of Direct Effect (InSIDE) assumption to be satisfied. The InSIDE assumption states that there is no correlation between the pleiotropic effects of genetic variants and the variant-exposure association. Additionally, the MR Egger method provides a test, known as the MR Egger intercept test, to detect directional pleiotropy. The deviation of the intercept from zero indicates evidence of directional pleiotropy (27).

Negative control outcome MR analysis was performed for the following outcomes: low hand grip strength (28) (48,596 cases; 207,927 controls; European ancestry), myopia (29) (36,623 cases; 419,031 controls; European ancestry), Parkinson disease (30) (15,056 cases, 18,618 proxy cases, 449,056 controls; European ancestry), and heel bone mineral density (31) (583,314 European ancestry individuals). These outcomes are selected as negative controls as they are currently not known to share strong pathological pathways with hypertension.

As BP summary data were obtained from a GWAS meta-analysis that adjusted for body mass index (BMI), collider bias might be introduced in the MR analysis. We performed a sensitivity analysis using BP summary data from a UKB GWAS that did not adjust for BMI (32).

To explore whether observed associations between antihypertensive drug classes and the outcomes are mediated through their BP-lowering effect, we performed MR analysis to investigate potential causal effects of genetically proxied SBP reduction on the outcomes of interest, leveraging genetic instruments throughout the whole genome.

LD clumping was performed using the TwoSampleMR package in R (33); IVW, weighted median, simple median, and MR Egger were performed using the MendelianRandomization package (version 0.6.0) in R (34).

### Summary-based Mendelian randomisation analysis

Summary-based Mendelian randomisation analysis (SMR) is a form of MR analysis that integrates GWAS and eQTL summary data to discover potential causal association of gene expression level with an outcome. Out of 49 tissues in the GTEx v8 (21), we performed the analysis on 29 tissues that are relevant to the pathophysiology of outcomes of interest and mechanism of the antihypertensive drugs. SMR estimate was presented as a beta coefficient with 95% CI per 1 unit increase in the expression of the gene in a particular tissue. To account for multiple SMR testing, a Bonferroni-corrected p-value of 0.0001 was used to explore significant associations; a p-value greater than 0.0001 but less than 0.05 indicates a nominal association.

HEIDI test is integrated into the SMR method to investigate if an observed association between gene expression and the outcome is due to a linkage scenario, in which the gene expression level and the outcome do not share a causal variant (14). A p_HEIDI_ of 0.01 was used as a threshold to indicate a linkage scenario - p_HEIDI_ < 0.01 indicates the observed association is likely due to a linkage scenario (14, 35).

SMR analyses and HEIDI tests were performed using SMR software version 1.3.1. Default settings were used: p_eQTL_ < 5×10^-8^, MAF > 0.01, and eQTL that are in very high or very low LD with the top associated eQTL (r^2^ > 0.9 or r^2^ < 0.05) were excluded from the HEIDI test to avoid collinearity (14).

### Colocalisation analysis

Colocalisation analysis was performed to investigate if an observed association between gene expression level and the outcome is due to a shared causal variant. We used coloc v5.2.1 R package to perform the analysis. Within a particular genomic region, a Bayesian test was performed to calculate posterior probabilities (PP) for five scenarios (15):

H_0_: There is no variant associated with either trait.

H_1_ and H_2_: There is a variant associated with only one of the two traits.

H_3_: There are distinct causal variants associated with each of the traits.

H_4_: There is a shared causal variant for both traits.

A high PP for H_4_ indicates a high probability of a shared causal variant for the traits. Whereas a high PP for H_3_ suggests that the observed association is likely due to a linkage scenario. We used default prior settings for the colocalisation analysis: p_1_ = 10^-4^, p_2_ = 10^-4^, p_12_ = 10^-5^.

## Results

### Genetic instruments

Throughout the entire genome, 454 uncorrelated SNPs (r^2^ < 0.001) were identified as genetic instruments for SBP (**Table S2)**. F-statistics for all of the SNPs were greater than 10, suggesting a low risk of weak instrument bias. Genetic instruments accounted for 4.7% of the total variance in SBP.

By using the predefined selection criteria, ten antihypertensive drug classes have genetic instruments for their effects (**Table S3**). These drug classes include AGTi (2 SNPs), ACEi (1 SNP), CCBs (29 SNPs), BB (7 SNPs), alpha-blocker (1 SNP), loop diuretic (4 SNPs), PSD (1 SNP), ERA (1 SNP), PDE5i (5 SNPs), and sGC stimulator (6 SNPs). F-statistics for all SNPs were greater than 10 (ranging from 30.48 to 352.48), which suggests that the risk of weak instrument bias is low.

### MR analysis of genetically proxied SBP reduction

Genetically predicted SBP reduction was significantly associated with a reduced risk of all the primary outcomes, with ORs for the outcomes ranging from 0.71 to 0.87 per 10 mmHg decrease in SBP (**Fig 2, Table S4**). The results obtained from the simple median, weighted median, and MR Egger methods were consistent with the primary MR analysis for all the outcomes except for T2D. The MR Egger intercept test indicated evidence of horizontal pleiotropy in the MR analysis of SBP with T2D risk (p-value intercept test = 0.0004; MR Egger OR per 10 mmHg decrease in SBP, 1.08, 95% CI, 0.92 - 1.26, p = 0.35) (**Table S4**).

**Figure 2-A.**
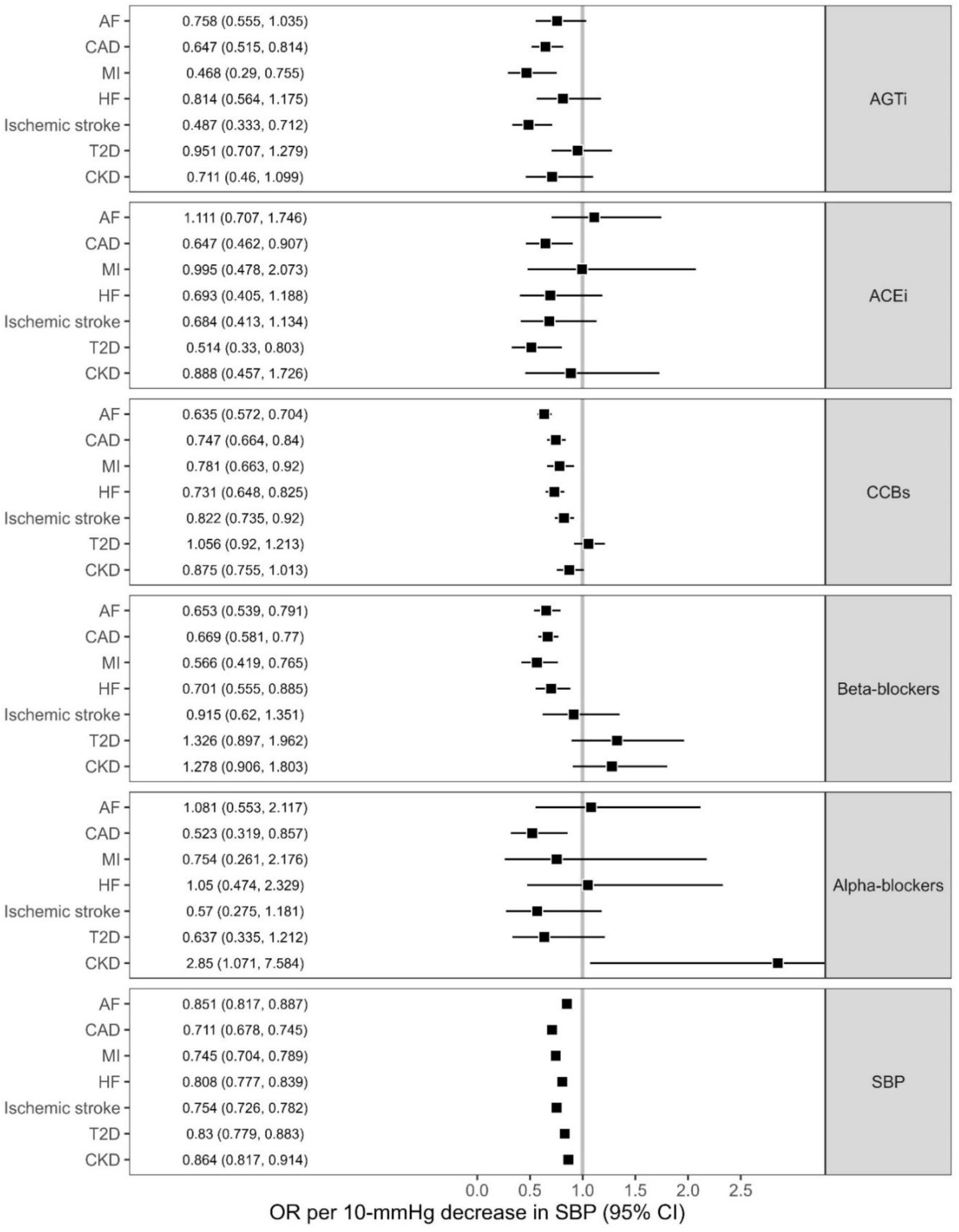
MR estimates for the effect of genetically lower blood pressure through the drug target genes of antihypertensive drug classes on the risk of the outcomes. Data were represented as odds ratio (OR) with 95% confidence interval (CI) of the outcome per 10 mmHg decrease in systolic blood pressure with risk for the outcomes of interest. OR of less than 1 suggests a decreased risk of disease associated with blood pressure lowering treatments, and vice versa. CAD indicates coronary artery disease; MI, myocardial infarction; AF, atrial fibrillation; HF, heart failure; CKD, chronic kidney disease; T2D, type 2 diabetes; AGTi, Angiotensinogen inhibitor; ACEi, angiotensin-converting enzyme inhibitors; CCBs, calcium channel blockers; SBP, systolic blood pressure.

**Figure 2-B.**
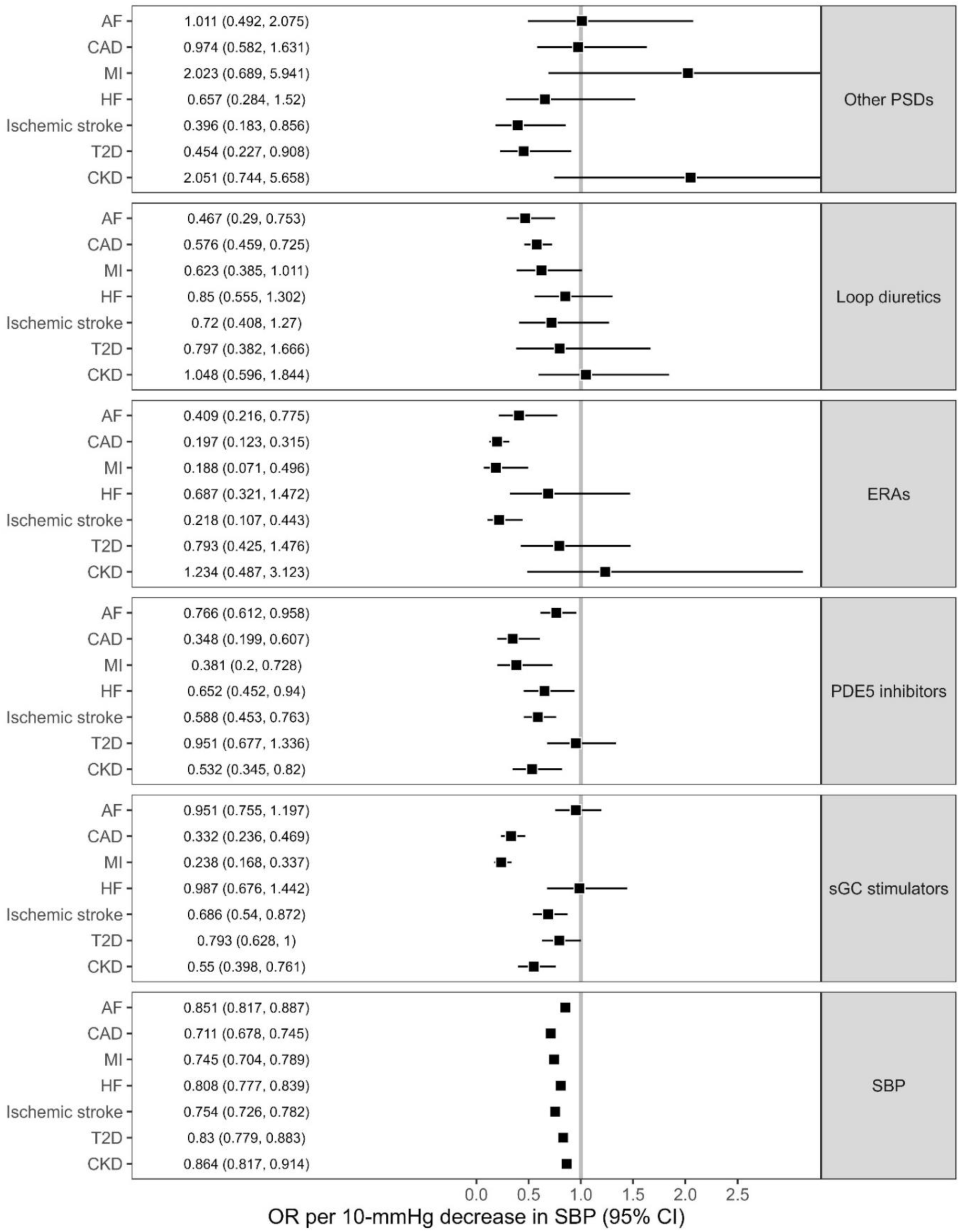
MR estimates for the effect of genetically lower blood pressure through the drug target genes of antihypertensive drug classes on the risk of the outcomes. Data were represented as effect size with 95% confidence interval (CI) of the outcome per 10 mmHg decrease in systolic blood pressure with risk for the outcomes of interest. FG, fasting glucose; AGTi, Angiotensinogen inhibitor; ACEi, angiotensin-converting enzyme inhibitors; CCBs, calcium channel blockers; Other PSDs, potassium-sparing diuretics (not include aldosterone antagonists); ERAs, endothelin receptor antagonists; PDE5 inhibitors, phosphodiesterase type 5; sGC; soluble guanylate cyclase; SBP, systolic blood pressure.

### MR analysis of antihypertensive drug classes

Genetically proxied antihypertensive drugs showed varying associations with cardiovascular outcomes (**Fig. 2-3, Table S5)**. Most drugs were associated with a reduced risk of CAD, with ACEi and alpha-blockers showing nominal association (OR ranging from 0.197 to 0.747 per 10 mmHg decrease in SBP). AGTi, CCB, ERA, and PDE5i showed protective effects against ischemic stroke, while there was suggestive evidence for sGC and PSD (OR range = 0.218 - 0.822). BB and sGC were associated with a reduced risk of MI; AGTi, CCB, ERA and PDE5i showed nominal associations (OR range = 0.187 - 0.781). For AF risk, we observed protective effects of CCB and BB, and suggestive evidence for loop diuretics, ERA, and PDE5i (OR range = 0.409 - 0.766). For HF, CCB showed a protective effect, while BB and PDE5i showed suggestive evidence of a protective effect (OR range = 0.652 - 0.731). sGC and PDE5i were associated with a reduced risk of CKD; whereas alpha-blockers showed a nominal association with an increased CKD risk. ACEi, sGC, and PSD were nominally associated with a reduced risk of T2D (OR range = 0.454 - 0.793). Results from the weighted median, simple median and MR Egger methods (where applicable) were consistent with those from the main MR analysis, with no horizontal pleiotropy detected in the MR Egger intercept test.

**Figure 3.**
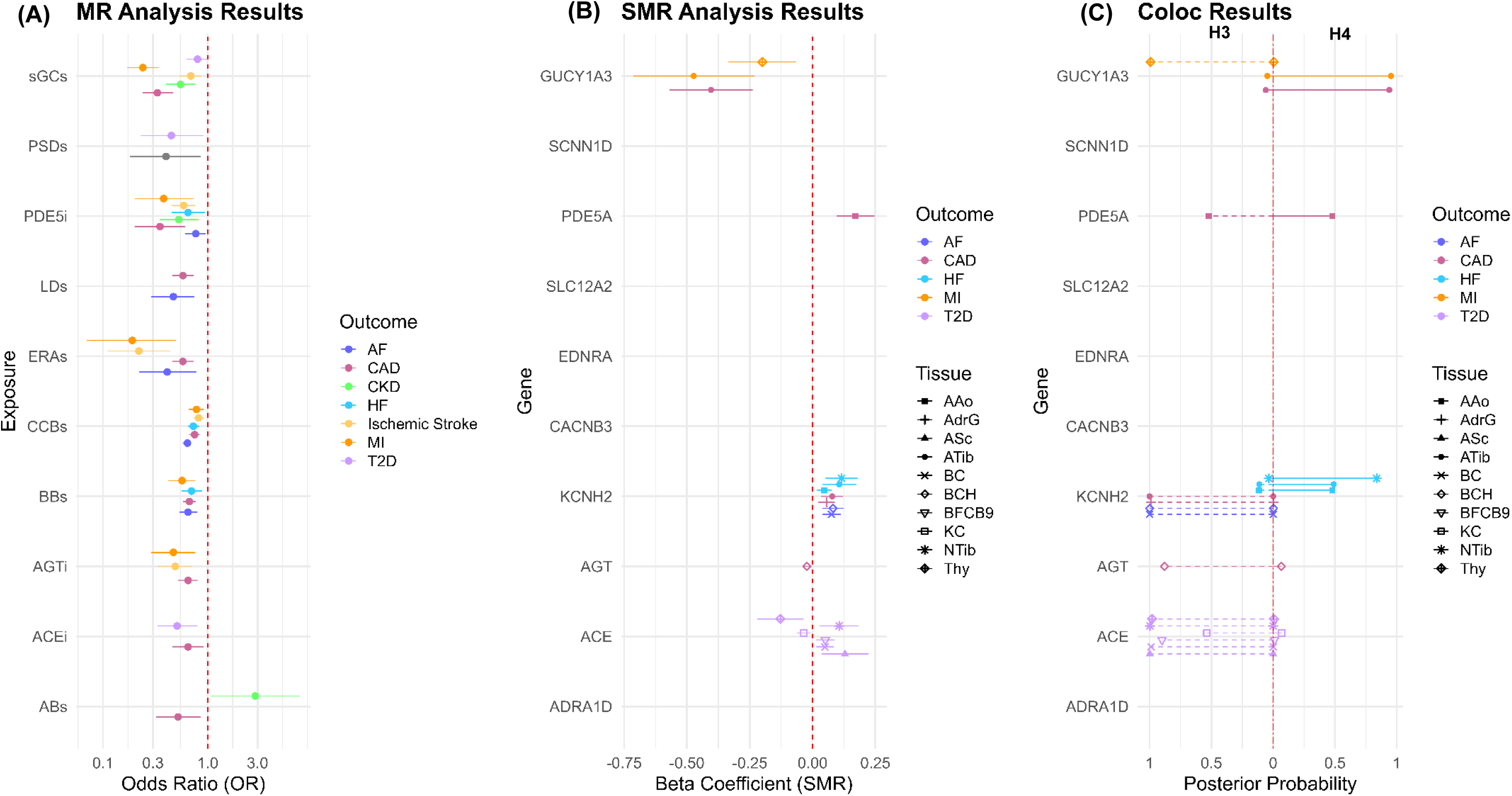
Results from MR, SMR, and colocalization analyses. Graph A shows MR results for drug classes that have association with the disease risk. Graph B shows SMR results for genes whose expression levels associated with the disease risk at p-value of 0.001; genes were placed at the same vertical point of the drug class that targets their proteins; full SMR results are shown in Fig 4, Fig S3-8. Graph C shows posterior probability for the scenario of distinct causal variants (H3, on the left of the vertical line) and of shared causal variant (H4, on the right). ASc, Adipose Subcutaneous; AdrG, Adrenal Gland; AAo, Artery aorta; Atib, Artery tibial; BCH, Brain Cerebellar Hemisphere; BC, Brain Cerebellum; BFCB9, Brain Frontal Cortex BA9; KC, Kidney Cortex; NTib, Nerve Tibial; Panc, Pancreas; Thy, Thyroid.

Negative control outcome analyses did not detect any significant association (**Table S6, Fig S1**). MR analysis using summary data for SBP obtained from a UKB GWAS gave consistent results with the primary MR analysis (**Table S7)**.

### SMR analysis of drug target gene expression

Out of 56 genes encoding the protein targets of the antihypertensive drugs, there were 19 genes whose expression levels were associated with SBP in at least one of the 21 tissues under the study (**Table S8, Fig S2**).

SMR was used to examine associations of gene expression levels with the disease outcomes across multiple tissues. Eighty-one associations were found across 15 tissues, which passed the HEIDI test. Three associations remain significant after accounting for multiple testing (p < 0.0001) (**Fig 3-4**, **Fig S3-8**. **Table S9-16)**. The expression levels of *GUCY1A3* (targeted by sGC stimulators) in the tibial artery and *PDE5A* (targeted by PDE5 inhibitors) in the aorta were significantly associated with CAD risk (p_SMR_ = 1.74×10^-06^, p_HEIDI_ = 0.345 for *GUCY1A3*, and p_SMR_ = 9.12×10^-06^, p_HEIDI_ = 0.338 for *PDE5A*). We also found a significant signal for the expression of the *KCNH2* gene (targeted by BBs) in the brain cerebellum with AF risk (p_SMR_ = 6.02×10^-05^, p_HEIDI_ = 0.195) (**Fig 3-4)**.

**Figure 4.**
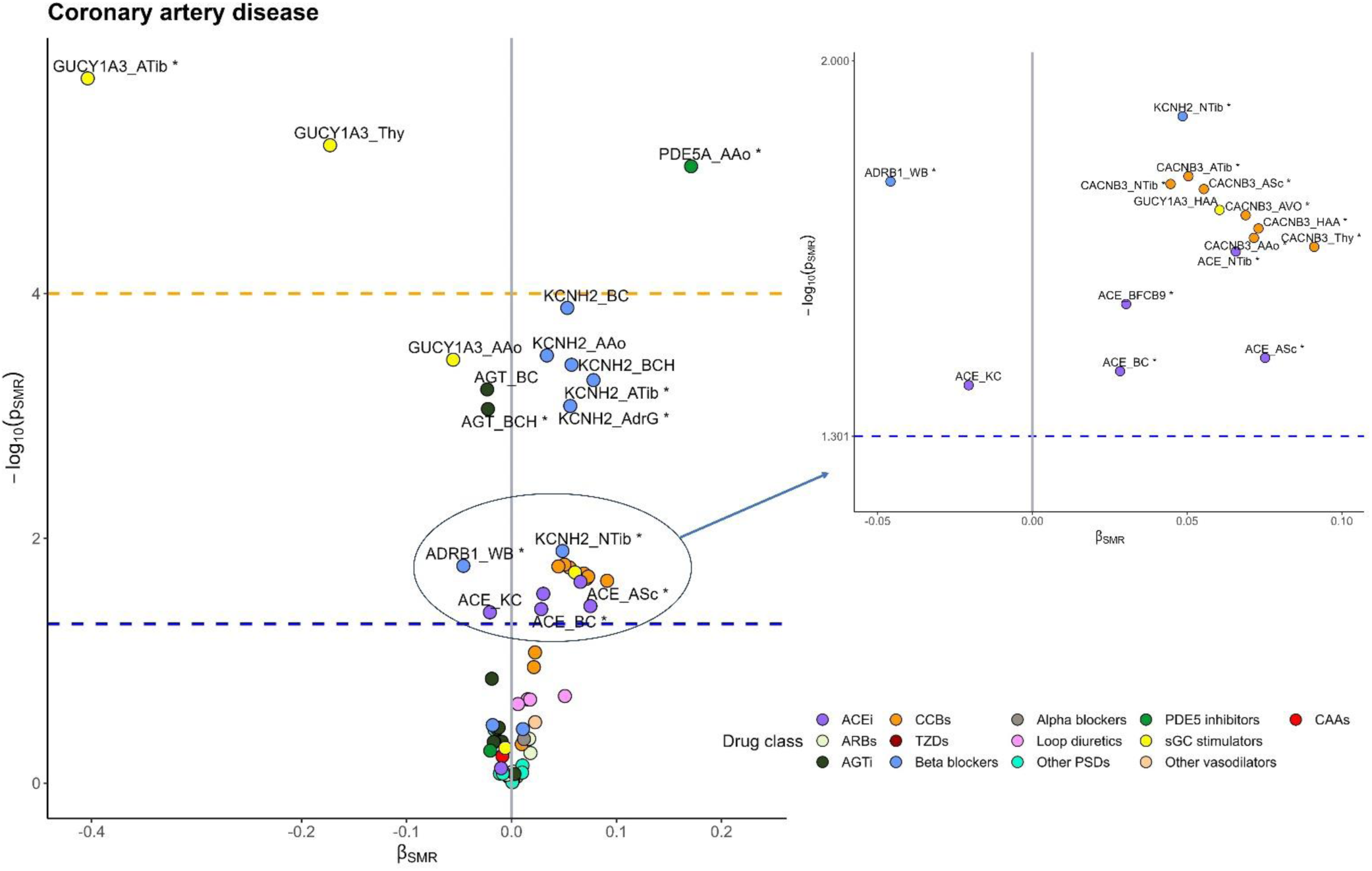
SMR associations of drug target gene-expression levels with CAD risk. The x-axis represents the β estimates from SMR analysis, and the y-axis represents the -log10(p_SMR_). The circle, which signifies the corresponding gene, was coloured according to the drug class that targets its protein. β_SMR_ were estimates from SMR analysis per 1 unit increase in the expression of the genes in a particular tissue. β_SMR_ < 0 suggests that an increase in gene expression levels in a particular tissue was associated with a decrease in CAD risk, and vice versa. The blue and orange dashed lines indicate a p-value of 0.05 and a Bonferroni-corrected p-value of 0.0001, respectively. The asterisk indicates HEIDI p-value greater than 0.01; The identifiers for each SMR association circle are the relevant gene name along with the following abbreviations for tissue types: AVO, Adipose Visceral Omentum; ASc, Adipose Subcutaneous; AdrG, Adrenal Gland; AAo, Artery aorta; Atib, Artery tibial, BCaud, Brain Caudate basal ganglia; BCH, Brain Cerebellar Hemisphere; BC, Brain Cerebellum; BCor, Brain Cortex; BFCB9, Brain Frontal Cortex BA9; BNacc, Brain Nucleus accumbens basal ganglia; BPut, Brain Putamen basal ganglia; HAA, Heart Atrial Appendage; HLV, Heart Left Ventricle, KC, Kidney Cortex, MSk, Muscle Skeletal, NTib, Nerve Tibial; Panc, Pancreas; Thy, Thyroid; WB, Whole Blood.

Colocalisation analysis gave a high posterior probability for a shared causal variant (H_4_ PP > 80%) for the following pairs: tibial artery *GUCY1A3* eQTL and CAD risk; tibial artery *GUCY1A3* eQTL and MI risk; and tibial nerve *KCNH2* and HF risk (**Fig 5, Table S11)**. For *PDE5A* expression in the aorta and CAD risk, the H_4_ posterior probability is 48%, whereas the H_3_ posterior probability is 52%. For *KCNH2* expression in the brain cerebellum and AF risk, we got a high H_3_ posterior probability (99%), suggesting that these traits have distinct causal variants in the gene region (**Table S11)**.

**Figure 5.**
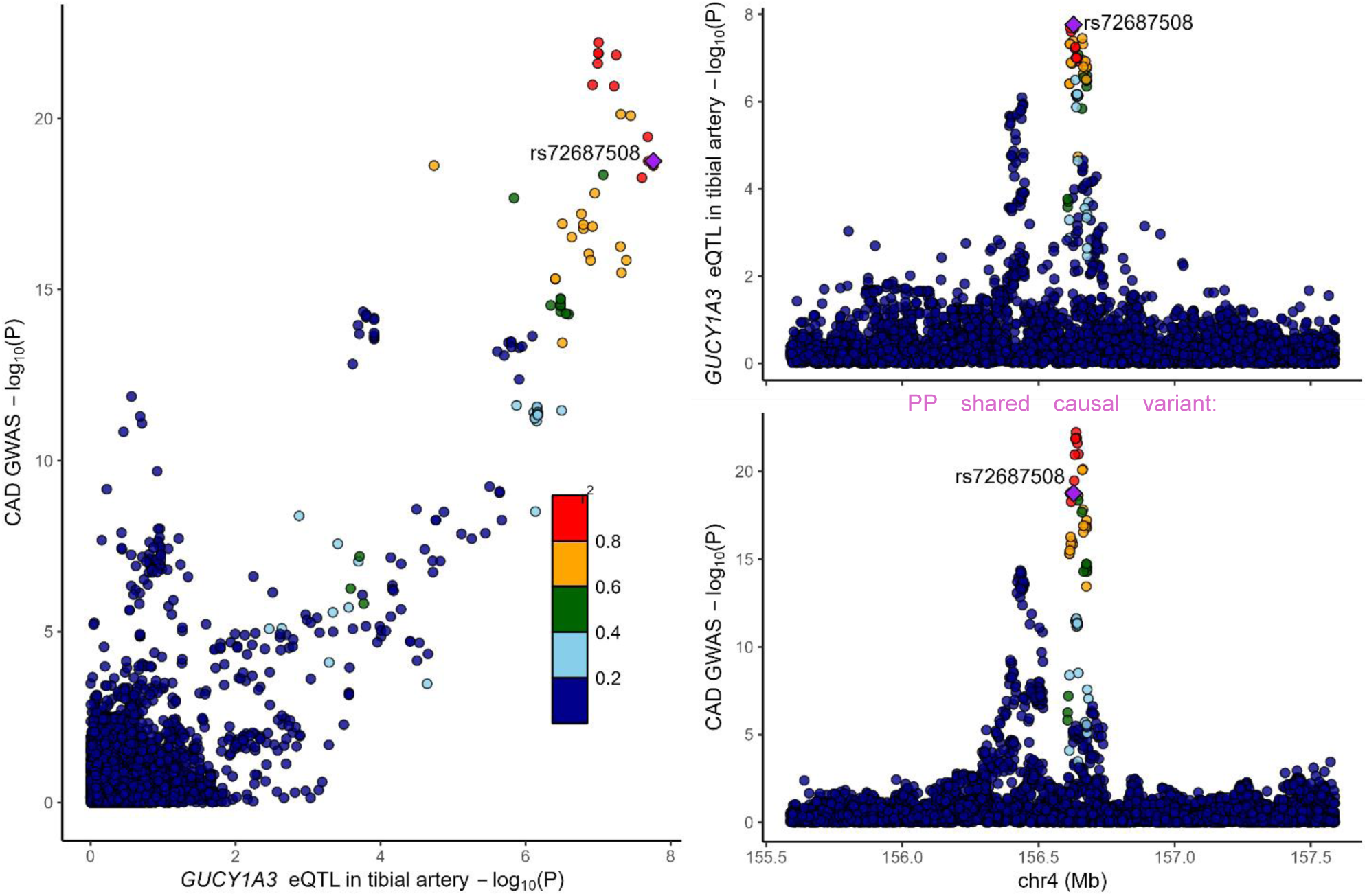
Colocalisation of eQTL in tibial artery for *GUCY1A3* and CAD risk. Colocalisation of genetic signal for *GUCY1A3* levels and CAD outcome in the 1-Mb window of the gene. SNPs are coloured according to their LD r^2^ with the lead SNP rs72687508.

## Discussion

Our study utilising an integrative approach that combines MR, SMR and colocalization analyses, provides evidence on the effectiveness of 10 antihypertensive drug classes across a range of cardiovascular, metabolic, and renal outcomes. While lower SBP is universally beneficial, with evidence of being causally associated with reduced risks of all studied outcomes, the effect of SBP lowering varied significantly depending on the antihypertensive drug class, revealing heterogeneity in their impact on different health outcomes.

CCBs demonstrated evidence of consistently protective effects across nearly all outcomes, except for T2D where no significant benefit was observed. BBs were evidenced to be particularly beneficial in reducing the risks of CAD, MI, AF and HF, though their effectiveness was inconclusive when considering other outcomes. AGTi were evidenced to be generally effective across most outcomes but had a null effect on T2D and did not reach statistical significance for HF and CKD. In contrast, ACEi presented mixed results, offering significant protection against CAD and T2D, but showing no effect on AF and MI, with inconclusive results for other outcomes. Alpha-blockers were associated with a protective effect against CAD but exhibited a detrimental impact on CKD. PSDs showed clear protective effects against stroke and T2D, while results for AF and CAD were null, and the effects on MI, HF, and CKD were inconclusive. Loop diuretics were protective against AF, CAD, and MI, had null effect on CKD and inconclusive for other outcomes. Among the newer or emerging therapies, ERAs were evidenced to show significant protective effects against AF, CAD, MI and stroke. PDE5 inhibitors were protective across all studied outcomes except T2D, sGC stimulators demonstrated protective effects against CAD, MI, stroke, T2D and CKD.

This study further integrates data from large-scale GWAS and eQTL studies to investigate whether the observed associations are mediated via gene transcription. We employed the SMR method, followed by the HEIDI test and colocalisation analysis to distinguish between linkage and true causality. The analyses were performed for 19 genes across 29 tissues that were broadly relevant to the pathophysiology of the outcomes and the mechanism of the antihypertensive drugs. SMR associations, which were corroborated by both HEIDI test and coloc, include *GUCY1A3* - CAD risk, *GUCY1A3* - MI risk in the tibial artery, and *KCNH2* - HF risk in the tibial nerve. We also found a significant association between *PDE5A* in the aorta and CAD risk, and between *KCNH2* in the cerebellum and AF risk, but coloc gave a low to moderate posterior probability for a shared causal variant.

Our selection criteria for genetic instruments for antihypertensive drug classes in two-sample MR is similar to the criteria applied in the previous study by Gill et al (19). Genetic variants were selected as SNPs located within or near the genes (which encode the protein targets of the drugs) and associated with SBP at p < 5×10^−8^. These SNPs were then clumped to a linkage disequilibrium threshold of r^2^ < 0.1 to ensure that the genetic instruments were uncorrelated. Furthermore, for SMR analysis we used significant eQTL for genes encoding the drug-target proteins as proxies for the effects of gene expression levels to evaluate whether the observed MR association is mediated through gene transcription. There is another approach to selecting genetic instruments for antihypertensive drugs, which was applied by several MR studies (36, 37). In this approach, genetic variants were selected as eQTL for the corresponding gene in any tissue using the GTEx v7. The selected SNPs were further validated by evaluating their association with SBP using a two-sample MR - SNPs that showed nominal association with SBP (p-value < 0.05) were retained in the main two-sample MR analysis. Using this approach, a recent MR study on atrial fibrillation found a preventative effect of SBP-lowering via alpha-blockers, beta-blockers, calcium channel blockers, and vasodilators (37). Our current MR study found significant protective effects of beta-blockers and calcium channel blockers and suggestive evidence for loop diuretics and vasodilators (ERAs and PDE5 inhibitors). Furthermore, our SMR analysis on AF risk found a significant signal for the expression of the *KCNH2* gene (targeted by BB) in the brain cerebellum, and nominal signals for this gene in the brain cerebellum hemisphere and tibial nerve.

Our study supports the effectiveness of sGC stimulators in reducing the risks of CAD, MI, and CKD. These findings align with previous research highlighting the role of the nitric oxide (NO)-sGC-cGMP pathway in cardiovascular protection (38–40).

The *GUCY1A3* gene, which encodes an alpha subunit of the sGC complex, has been linked to CAD and MI in large-scale GWAS (38, 39). Our results show that increased *GUCY1A3* expression in the tibial artery is associated with lower SBP and a reduced risk of CAD and MI, reinforcing the potential of sGC stimulation as a therapeutic strategy for CAD management.

PDE5 inhibitors, commonly used for erectile dysfunction and pulmonary hypertension, were also found to have cardioprotective effects (41, 42). Biological plausibility is supported by its known function through inhibition of the breakdown of cGMP, thereby enhancing NO activity, which has been associated with cardiovascular benefits. Our MR study supports this, showing a causal association between PDE5 inhibition and reduced risks of CAD and MI. This was corroborated by SMR findings, which linked increased *PDE5A* expression in the aorta to higher SBP and increased CAD risk, suggesting a potential role for PDE5 inhibitors in cardiovascular disease management.

We also observed suggestive associations between *KCNH2* gene expression in the tibial nerve and HF risk, with colocalisation analysis indicating a shared causal variant. The *KCNH2* gene, known for its role in generating repolarising potassium currents, has been implicated in long QT syndrome and drug-induced QT prolongation, conditions associated with an increased risk of sudden cardiac death (43). Our findings further support the association of beta-blockers with reduced risks of HF and AF in relation to hypertension, highlighting KCNH2 as a potential target for therapeutic intervention.

Our study is the first study to conduct this in-depth investigation of the effects of a wide range of antihypertensive drug classes - at the drug class level as well as the individual target gene expression level - on a broad spectrum of diseases, including cardiovascular, diabetic and renal diseases. The integration of MR, SMR, and colocalization analyses allowed us to triangulate evidence and enhance the credibility of our findings. The convergence of MR and SMR results for PDE5 inhibition, sGC stimulation, and ACE inhibition underscores the robustness of these findings. By corroborating results across different analytical methods, we reduced the likelihood of bias and confounding, thereby strengthening the causal inferences.

Our study has limitations. Firstly, as MR estimates the cumulative effect of lifetime exposure to genetic variants, they may differ from the effects obtained from a clinical intervention such as an antihypertensive treatment. Although the discrepancy suggests that MR estimates may not precisely reflect the effect size of pharmacological intervention, MR still serves as a valuable tool to detect the existence and directionality of causal effects. Secondly, some of the drug classes do not have genetic instruments, thus, the study was unable to investigate the potential causal effects of those. Future studies that include a GWAS with a large sample size for blood pressure (44) could potentially identify such variants and therefore could provide a more profound understanding of the varying effects of different antihypertensive drug classes. Thirdly, tissue misspecification is another limitation of the study, given the limited availability of eQTL data for some specific tissues. eQTL studies with sufficient sample sizes will help alleviate this issue. Finally, summary data used in this study was restricted to individuals of European descent, potentially limiting the generalizability of the results to this specific group. The availability of large-scale GWAS datasets from more ethnically diverse populations will allow future MR studies to investigate potential ethnic differences.

## Conclusion

Our findings demonstrate the power of integrating MR, SMR, and colocalization analyses to uncover the potential differential effects of various antihypertensive drug classes on a broad spectrum of cardiometabolic and renal outcomes. The heterogeneity observed in drug efficacy across different outcomes underscores the potential of personalised antihypertensive therapy. These results not only validate the clinical relevance of existing antihypertensive therapies but also highlight new therapeutic targets, such as *GUCY1A3* and *PDE5A*, for further exploration. Future research should focus on validating these findings in diverse populations and exploring the mechanistic pathways that link blood pressure regulation to various health outcomes, ultimately paving the way for more personalized and effective hypertension management strategies.

## Data Availability

All data referred to in this study are available upon reasonable request to the authors

## Acknowledgement

Conceptualization: S.P.; Data curation: N.N.L.; Formal analysis: N.N.L.; Methodology: N.N.L., T.Q.B., D.G. and S.P.; Supervision: J.M., D.G. and S.P.; Writing – original draft: N.N.L.; Writing - review & editing: N.N.L., T.Q.B., J.M., D.G. and S.P.

